# “Learn from the lessons and don’t forget them”: Identifying transferable lessons for COVID-19 from meningitis A, yellow fever, and Ebola virus disease vaccination campaigns

**DOI:** 10.1101/2021.07.15.21260439

**Authors:** Julie Collins, Rosie Westerveld, Kate A Nelson, Hana Rohan, Hilary Bower, Siobhan Lazenby, Gloria Ikilezi, Rebecca Bartlein, Daniel G Bausch, David S Kennedy

## Abstract

**Introduction:** COVID-19 vaccines are now being distributed to low- and middle-income countries (LMICs), with global urgency surrounding national vaccination plans. LMICs have significant experience implementing vaccination campaigns to respond to epidemic threats but are often hindered by chronic health system challenges. We sought to identify transferable lessons for COVID-19 vaccination from the rollout of three vaccines that targeted adult groups in Africa and South America: MenAfriVac (meningitis A); 17D (yellow fever); and rVSV-ZEBOV (Ebola virus disease).

**Methods:** We conducted a rapid literature review and 24 semi-structured interviews with technical experts who had direct implementation experience with the selected vaccines in Africa and South America. We identified barriers, enablers, and key lessons from the literature and from participants’ experiences. Interview data were analysed thematically according to seven implementation domains.

**Results:** Participants highlighted multiple components of vaccination campaigns that are instrumental for achieving high coverage. Community engagement is an essential and effective tool, requiring dedicated time, funding and workforce. Involving local health workers is a key enabler, as is collaborating with community leaders to map social groups and tailor vaccination strategies to their needs. Vaccination team recruitment and training strategies need to be enhanced to support vaccination campaigns. Although recognised as challenging, integrating vaccination campaigns with other routine health services can be highly beneficial if well planned and coordinated across health programmes and with communities.

**Conclusion:** As supplies of COVID-19 vaccines become available to LMICs, countries need to prepare to efficiently roll out the vaccine, encourage uptake among eligible groups, and respond to potential community concerns. Lessons from the implementation of these three vaccines that targeted adults in LMICs can be used to inform best practice for COVID-19 and other epidemic vaccination campaigns.

KEY QUESTIONS

What is already known?

- Low- and middle-income countries (LMICs) have substantial experience conducting vaccination campaigns as a part of epidemic responses.
- Vaccination campaigns in LMICs are impacted by a number of systemic challenges, including poor infrastructure, limited resources, and an overstretched health workforce.
- Meningitis A, yellow fever and Ebola virus disease vaccines have been recently rolled out in LMICs to respond to epidemic threats. These campaigns share some of the same challenges anticipated for COVID-19 vaccination, including the focus on adult target groups.

What are the new findings?

- Extensive community engagement is crucial when targeting adults for vaccination in LMICs to shift community perceptions that vaccination is only associated with children.
- Working with community leaders to map social groups and plan effective vaccination strategies is vital to achieving high vaccination coverage.
- Recruiting local health workers who have established links to the community, can speak the local language, and can leverage existing rapport to increase vaccination uptake, is preferred over bringing in staff from other regions.
- Vaccination training quality is reduced as information is transmitted down to lower levels using the ‘cascade’ or ‘training-of-trainers’ model. Training for vaccination campaigns in LMICs has been further affected by COVID-19 and the move to remote learning. Where access to training is limited, a greater emphasis is placed on resource-intensive supervision to ensure the effectiveness of vaccination campaigns.

What do the new findings imply?

- Previous vaccination campaigns conducted during epidemics are an important source of transferable lessons that can assist countries in their COVID-19 vaccine rollouts and future epidemic preparedness.
- Our findings suggest that countries can strengthen vaccination campaigns during epidemics by recruiting local health workers to assist vaccination teams, by providing operational funding for pre-campaign community engagement and social mobilisation activities, and by examining the effectiveness of vaccination training and developing new models where needed.
- Implementing these lessons for COVID-19, however, relies on countries having sufficient vaccine supply.

## INTRODUCTION

Development and mass-production of multiple safe and effective COVID-19 vaccines have progressed faster than expected. There are signs that, with high population-level vaccination coverage, pre-pandemic levels of mobility and economic activity could safely resume.[1, 2] However, COVID-19 vaccination has been dominated by high-income countries, which have purchased 51% of vaccine doses yet represent only 14% of the global population.[3] As of 10 July 2021, only 1% of people in low-income countries had received at least one dose of a COVID-19 vaccine.[4] Adequate vaccine supply is a critical first step. Access mechanisms, such as the COVID-19 Vaccines Global Access (COVAX) initiative, have been established to support vaccine supply in low- and middle-income countries (LMICs); however, modest initial targets have not been met.[5] In addition to increasing access to vaccine supplies, critical work is needed to ensure that vaccines reach their intended target groups.

Many factors can complicate vaccine rollouts in LMICs, including limited health and surveillance infrastructure, insufficient cold chain transportation and storage capacity, and an under-resourced health workforce.[6, 7] However, some LMICs also have substantial experience in conducting vaccination campaigns to prevent or respond to epidemics, from which valuable lessons can be drawn for the rollout of COVID-19 vaccines.

We examined vaccination campaigns recently conducted as part of epidemic responses (hereafter, ‘vaccination campaigns’) in LMICs in Africa and South America. We selected three vaccines – MenAfriVac (meningitis A), 17D (yellow fever), and rVSV-ZEBOV (Ebola virus disease) – that share some of the same challenges anticipated for COVID-19 in LMICs (Table 1) to capture insights and make recommendations for countries implementing COVID-19 vaccination campaigns.

**TABLE 1:** Characteristics of the selected vaccines.

| **Criteria** | **Ebola virus disease**<br>**(rVSV-ZEBOV)** | **Yellow Fever**<br>**(17D)** | **Meningitis A**<br>**(MenAfriVac)** |
| --- | --- | --- | --- |
| **Cold chain requirements** | -70°C | 2-8°C [61] | 2-8°C[62] |
| **Dose regimen** | Single Dose | Single Dose | Single Dose[63] |
| **Target populations during outbreak-related campaigns** | Adults aged ≥18 years.*<br>Healthcare workers, contacts of confirmed cases & contacts of contacts. | Children and adults† aged ≥9 months[61] | Children and adults aged 1 – 29 years[63] |
| **Regions** | Democratic Republic of Congo, Burundi, Guinea, Liberia, Sierra Leone, Rwanda, Uganda | Africa and South America | "Meningitis belt" in Africa – sub-Saharan Africa from Senegal to Ethiopia [62] |
| **Year developed/licenced** | 2019[64] | 1927[61] | 2010[62] |
| **Key similarities with COVID-19 Vaccines** | Primarily targeted adults; required ultra-cold chain management | Targeted adults as well as children | Targeted adults as well as children |

\*Children aged 6–17 years also included in some Phase 1 to 3 trials[65]; Children >6 months were included in the 2018–2020 outbreak in Democratic Republic of Congo under compassionate use protocol.[66]
†Upper limit age group varied by outbreak.

## METHODS

We conducted a rapid literature review followed by semi-structured interviews with technical experts to identify barriers, enablers, and lessons from implementing the three vaccines in recent outbreaks in Africa and South America.

We designed a thematic framework of seven domains (Table 2) to guide the research, based on existing vaccine readiness assessment tools and gap analyses developed for implementing Ebola virus disease (EVD) vaccines[8, 9] and COVID-19 vaccination guidelines developed by the World Health Organization (WHO).[10]

**TABLE 2:** Thematic framework for the implementation of vaccines.

| Domain | Description |
| --- | --- |
| 1) Planning and coordination | Macro/microplanning; funding; vaccination campaign management; decision-making; coordination and communication mechanisms; stakeholder engagement; stakeholder roles; policy and regulatory framework. |
| 2) Target groups and delivery strategies | Target group inclusion and exclusion criteria; prioritisation of groups; communication of target groups; delivery strategies for vaccination (e.g., house-to-house, fixed post, mobile fixed post); tailoring of delivery strategies to reach sub-groups. |
| 3) Logistics and supply | Supply chain; infrastructure; vaccine storage; cold chain (and ultra-cold chain) management; transportation; equipment (including personal protective equipment); waste disposal. |
| 4) Vaccination teams | Team composition and roles; recruitment; training techniques and processes; team coordination and communication. |
| 5) Vaccination monitoring and safety surveillance | Identifying cases of disease and differentiating between similar pathogens; recording, reporting and monitoring vaccination coverage; identification, reporting and management of adverse events following immunisation; use of technology. |
| 6) Community engagement and social mobilisation<sup>1</sup> | Developing relationships with communities and working together to conduct vaccination activities; strategies to increase vaccine demand and uptake, including communication strategies. |
| 7) Vaccine confidence<sup>2</sup> | Perceptions and attitudes toward the vaccine; factors contributing to confidence or resistance; types of rumours and misinformation; strategies to address rumours and misinformation. |

<sup>1</sup>World Health Organization definitions of community engagement and social mobilisation were used in this research, however it was noted that these terms were often used interchangeably by key informants.[67, 68]
<sup>2</sup>Vaccine confidence was not identified in articles retrieved in the literature review but was discussed in the key informant interviews.

### Literature review

We searched Embase, MEDLINE & Global Health databases in early February 2021. Searches were not restricted by location, year of publication or language. Search terms are provided in Appendix 1. For inclusion, articles were required to 1) focus on an LMIC in Africa or South America, and 2) describe the implementation of one of the selected vaccines. Following de-duplication, we conducted title and abstract screening, followed by full-text screening. Texts meeting these criteria were cross-referenced to identify additional relevant papers for inclusion. Data were extracted from the included articles according to the thematic framework (Table 2).

### Key informant interviews

#### Participants

Using a purposive sampling approach, we identified technical experts through the professional networks of the research team and via “snowball sampling.” Participants had either 1) involvement in the rollout of one or more of the selected vaccines through global initiatives or organisations, or 2) implementation experience with one or more of the selected vaccines at either a regional, national, or sub-national level.

#### Data collection

We developed a semi-structured interview guide, based on preliminary findings from the literature review and using the thematic framework (Table 2). The interview guide helped elicit specific information across the domains but was sufficiently flexible to allow the interview to be guided by the participants’ experiences. While the interviews focused on the three selected vaccines, interviewers did not dissuade participants from drawing on their experiences from other campaigns. Participants were given the opportunity to reflect on their experiences with the initial COVID-19 vaccination rollout, where appropriate.

Interviews were conducted virtually via Zoom and were in either English or French. Interviews were digitally recorded, professionally translated to English when required, and transcribed. The interviewers verified each transcript to ensure accuracy before commencing data analysis.

#### Data analysis

We analysed the qualitative data thematically, using deductive and inductive coding in Nvivo 12 Plus. A coding framework was developed based on the seven domains in the thematic framework (Table 2), along with additional themes emerging from the data. Key trends, barriers, enablers, and lessons shared by participants were identified across each theme.

## RESULTS

### Literature review

Thirty-seven studies met the inclusion criteria (Figure 1): 15 meningitis A[11–25]; 11 Ebola virus disease[26–36]; eight yellow fever[37–44]; one meningitis A and yellow fever[45]; and two focusing on general vaccination strategies.[46, 47] Twenty-eight studies focused on African countries[11–35, 40, 44, 46, 47]; two on South American countries[41, 43]; and six studies did not specify a region[36–39, 42, 45] (Table 3).

**TABLE 3:** Studies identified through the literature review.

| Author | Year | Disease |  |  |  | Location |  |  |
| --- | --- | --- | --- | --- | --- | --- | --- | --- |
|  |  | Meningitis A | Ebola | Yellow fever | General | Africa | South America | Global |
| *Aguado, M.T., et al.[11]* | 2015 | X |  |  |  | X |  |  |
| *Burchett, H.E.D., et al.[12]* | 2014 | X |  |  |  | X |  |  |
| *Marchetti, E., et al.[13]* | 2012 | X |  |  |  | X |  |  |
| *Zipursky, S., et al.[14]* | 2014 | X |  |  |  | X |  |  |
| *Djingarey, M.H., et al.[15]* | 2012 | X |  |  |  | X |  |  |
| *Cibrelus, L., et al.[16]* | 2015 | X |  |  |  | X |  |  |
| *Okwo-Bele, J.M. et al.[17]* | 2011 | X |  |  |  | X |  |  |
| *Tartof, S., et al.[18]* | 2013 | X |  |  |  | X |  |  |
| *Mbaeyi, S., et al.[19]* | 2020 | X |  |  |  | X |  |  |
| *Daugla, D.M., et al.[20]* | 2014 | X |  |  |  | X |  |  |
| *Nkwenkeu, S.F., et al.[21]* | 2020 | X |  |  |  | X |  |  |
| *Patel, J.C., et al.[22]* | 2019 | X |  |  |  | X |  |  |
| *Diomande, F.V.K., et al.[23]* | 2015 | X |  |  |  | X |  |  |
| *Berlier, M., et al.[24]* | 2015 | X |  |  |  | X |  |  |
| *Djingarey et al.[25]* | 2015 | X |  |  |  | X |  |  |
| *Ughasoro, M.D., et al.[26]* | 2015 |  | X |  |  | X |  |  |
| *Wolf, J., et al.[27]* | 2020 |  | X |  |  | X |  |  |
| *Jusu, M.O., et al.[28]* | 2018 |  | X |  |  | X |  |  |
| *Samai, M., et al.[29]* | 2018 |  | X |  |  | X |  |  |
| *Dean, N.E., et al.[30]* | 2019 |  | X |  |  | X |  |  |
| *Hossmann, S., et al.[31]* | 2019 |  | X |  |  | X |  |  |
| *Grantz, K.H., et al.[32]* | 2019 |  | X |  |  | X |  |  |
| *Juan-Giner, A., et al.[33]* | 2019 |  | X |  |  | X |  |  |
| *Elemuwa, C., et al.[34]* | 2015 |  | X |  |  | X |  |  |
| *Alenichev, A. et al.[35]* | 2020 |  | X |  |  | X |  |  |
| *Folayan, M.O., et al.[36]* | 2016 |  | X |  |  |  |  | X |
| *Tomashek, K., et al.[37]* | 2019 |  |  | X |  |  |  | X |
| *Chen, L.H. et al.[38]* | 2017 |  |  | X |  |  |  | X |
| *Vannice, K. et al.[39]* | 2018 |  |  | X |  |  |  | X |
| *Barrett, A.D.T. et al.[40]* | 2016 |  |  | X |  | X |  |  |
| *Possas, C., et al.[41]* | 2018 |  |  | X |  |  | X |  |
| *Martins, R.M., et al.[42]* | 2013 |  |  | X |  |  |  | X |
| *Flamand, C., et al.[43]* | 2019 |  |  | X |  |  | X |  |
| *Legesse, M., et al.[44]* | 2018 |  |  | X |  | X |  |  |
| *Nguyen, T. et al.[45]* | 2019 | X |  | X |  |  |  | X |
| *Yakum, M.N., et al.[46]* | 2015 |  |  |  | X | X |  |  |
| *Sow, C., et al.[47]* | 2018 |  |  |  | X | X |  |  |

**FIGURE 1:**
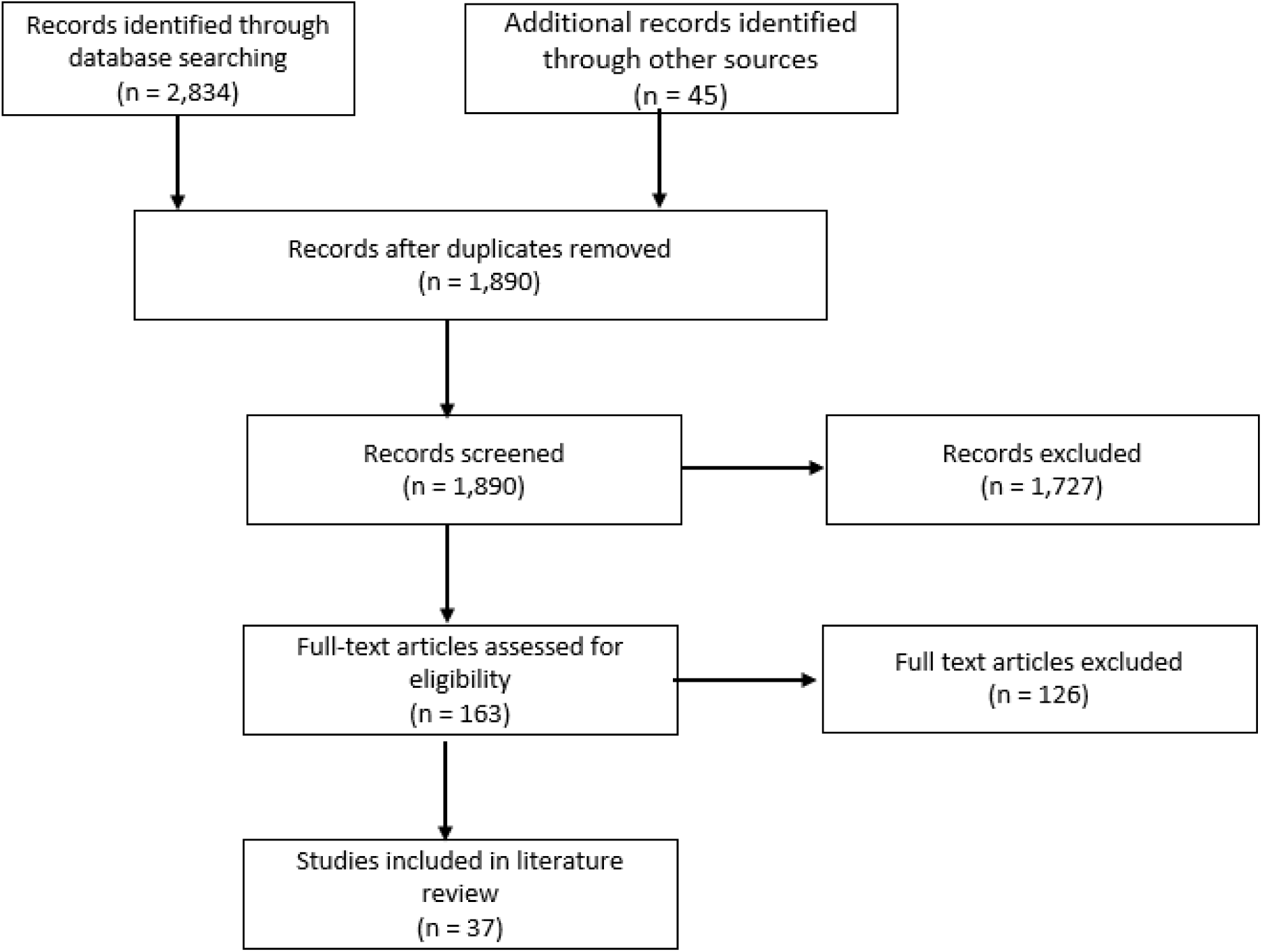
Literature review flowchart.

### Key informant interviews

Twenty-four key informants were interviewed from late February to late April 2021. Participants included health professionals and technical officers working for national health agencies, multilateral organisations (including United Nations agencies), non-governmental organisations (NGOs), and academia. All participants had worked with at least one of the selected vaccines in LMICs, with 23 reflecting on their vaccination campaign experiences across West, Central and East Africa, and one on their experiences in South America. Sixty-three per cent of participants were male (15/24). The average interview length was one hour (range 40–80 minutes).

### Key lessons for COVID-19 vaccination

We present findings from the literature review and qualitative interviews together, according to the thematic framework. Illustrative quotes have been included along with the participant’s organisation type and regions where they worked with the selected vaccines.

#### 1) Planning and coordination

##### Early engagement with diverse stakeholders to support campaign planning

Successful vaccination campaigns engage early with a broad network of stakeholders to support detailed pre-campaign planning. Collaboration with other government departments (e.g., Ministries of Education, Finance, and Transport) was considered crucial by interview participants, since health teams often relied on their support to conduct activities. Involving staff at the health facility level was critical to understand context, geography and social nuances within their catchment area.

> For the success of the campaign, we must have a strong microplan, originating from the health facility itself that is going to be involved. … [without this] you may still succeed in the vaccination coverage, but you will have a lot of hurdles. (National health agency, East Africa)

While participants noted the time pressures of reactive vaccination campaigns, the importance of engaging and planning with health facility staff and communities before commencing vaccination was consistently emphasised.

International partners, such as multilateral organisations, NGOs, and civil society organisations, play a key role in supporting campaign resourcing and implementation. Disease-specific initiatives, such as the Meningitis Vaccine Project and the Eliminate Yellow Fever Epidemics strategy, have assisted with access to vaccines, supported vaccine implementation and promoted mutual learning between countries.[13] However, participants did raise concerns about the sustainability of mass vaccination activities without such external support.[23–25]

COVAX and other initiatives are working to improve access to COVID-19 vaccines for LMICs. Still, some participants called attention to a gap in operational funding needed to support vaccination activities, such as community engagement.

> As long as the funds for operations are available, then it will be much easier for us to roll out any vaccine activity. … Fine, you are giving us the vaccine free of charge, but you should also factor in the operational costs. (Multilateral organisation, East Africa)

Insufficient operational funding has impaired some countries’ capacity to conduct preparatory activities for COVID-19 vaccination campaigns, such as raising awareness and encouraging vaccine uptake among health workers; participants were apprehensive about vaccine uptake in the absence of such activities.

##### Establishing strong coordination mechanisms

Robust coordination mechanisms are necessary to provide direction and oversight over campaign activities. Participants highlighted the benefits of an incident management system (IMS) that delineates the roles and responsibilities of government departments (e.g., disease control, routine immunisation, national drug authorities) and partner organisations and prevents the duplication of response efforts.

> [The IMS] helped us to coordinate the partners, because at the beginning of Ebola in West Africa, everybody took the money, went into the community without asking the others. [It was a] nightmare, until we found this coordination mechanism, this harmonisation of all the priorities in one single strategic response plan. (Multilateral organisation, West and Central Africa)

Within the IMS, technical working groups (e.g., logistics, social mobilisation) can foster collaboration between health agencies and international partners. In relation to COVID-19, some participants outlined how they were drawing on the strength of previously established technical working groups to conduct rapid vaccination preparations. The need for effective communication both within and between working groups was emphasised during interviews. Regarding EVD vaccination campaigns, some participants highlighted how the ‘industry’ of the outbreak led to competition and fragmentation. Different response teams (e.g., contact tracing and vaccination teams) vied to demonstrate their unique contributions and retain high-paying positions.[32]

#### 2) Target groups and delivery strategies

##### Target groups and community perceptions

Vaccinating adults requires a shift in community understandings of vaccination, which are traditionally associated with children. Perceptions and suspicions around adult target groups need to be recognised and accounted for in vaccination planning. Participants highlighted how communities might have concerns around both being included and excluded from vaccination target groups. For example, the inclusion of adults of reproductive age was frequently associated with concerns that vaccines affected fertility. On the other hand, some participants noted concerns from pregnant women about being initially excluded from receiving the rVSV-ZEBOV vaccine. Younger adults, notably young males, were described in both the literature and interviews as often being indifferent to meningitis A or yellow fever vaccination; their perceived risk of contracting the disease or developing severe outcomes was low so they did not see the benefit of being vaccinated. Interview participants also outlined how older adults sometimes needed convincing that it was worthwhile to vaccinate them in their stage of life.

> “No, no, no, at my age,” some will tell you, “at my age I think I’m just about due, [to die].” … [We use] persuasions in line with trying to make them valuable to society. (National health agency and academia, East Africa)

Participants spoke of spending considerable time explaining the disease and the vaccine development, safety, and regulatory processes to communities, along with why adults were being targeted for vaccination. This type of meaningful engagement with communities is critical to facilitate a better understanding of the disease and address community concerns around vaccination target groups. However, participants noted that time and resources (e.g., staff, development of messages, training, transport, and per diems) are required for this process.

##### Tailoring delivery strategies

Tailored delivery strategies are required for groups that experience either physical or social barriers to accessing vaccination. For example, young adult males were sometimes unable to access vaccination sites due to work commitments during meningitis A campaigns. To improve uptake, vaccination centres were positioned near facilities they frequented, such as workplaces, hotels, restaurants, and transport hubs.[25] Social and cultural norms were also identified as potential barriers to vaccination. During meningitis A campaigns in West Africa, sociocultural beliefs sometimes prevented men from attending vaccination centres at the same time as women and children.[25] Similarly, one participant (Multilateral organisation, East and West Africa) described how young married women might not be able to attend a public vaccination site without authorisation from their spouses. Engaging with community leaders to map social groups and identify when, where, and how vaccination teams can most effectively reach sub-groups is vital to achieving high vaccination coverage.

##### Integrating vaccination campaigns with other health services

Participants highlighted how accessing remote locations during vaccination campaigns can be viewed as an important opportunity to provide other health services. Integrating multiple vaccination campaigns can minimise time burdens on both communities and health workers and can potentially increase uptake by offering additional health services that the community prioritises. For example, one participant (National health agency, West Africa) described how vaccination uptake in nomadic communities increased following the development of a collaborative human-animal health initiative where vaccinations and health assessments for community members and their livestock were conducted during the same visit. While some participants viewed multi-service integration positively, they also mentioned challenges around the coordination and reporting of multiple activities.

Integrating COVID-19 vaccination with other services requires careful planning with communities to prevent potential spillover of COVID-19 vaccine hesitancy to other services. In some settings, participants had already observed an adverse effect on the uptake of other vaccines due to COVID-19.

> Some [community members] are really against COVID-19 vaccination, they say “no, you want to smuggle in the COVID-19 vaccine in the name of yellow fever.” (National health agency, West Africa)

Consequently, participants outlined how vaccination teams have had to differentiate between vaccines for COVID-19 and vaccines for other pathogens, reassuring communities that the dispensed vaccine was *not* a COVID-19 vaccine. This differentiation was seen to be the only way to ensure that the uptake of other vaccines was not compromised, but it adds to the complexity of messaging for COVID-19 vaccines.

#### 3) Logistics and supply

##### Vaccine storage and transportation

Participants outlined how vaccine storage assessments were necessary before each campaign and stressed how quickly capacity could change at the facility level, affecting product viability. During meningitis A clinical trials, mock-up shipments of MenAfriVac were sent to each country to test the supply chain and ensure the vaccines had been correctly handled throughout their journey.[13] In-depth logistical planning exercises and simulations can help identify and mitigate potential areas of vaccine wastage.[12, 48]

Subnational vaccine storage hubs that enable the rapid movement of supplies during a campaign are critical to success, particularly in areas where it is difficult to estimate the target population. Some vaccines (i.e., rVSV-ZEBOV, mRNA vaccines for COVID-19) require ultra-cold chain (UCC) infrastructure. While centralised depots with reliable access to electricity may seem most effective, some participants felt that centralisation led to unacceptable delays. Increasing storage capacity and cold chain management at the subnational level was seen as a solution to ensure the timely distribution of vaccines during a campaign.

During EVD vaccine clinical trials, significant investments were made to ensure appropriate UCC infrastructure was in place.[28] Interview participants believed that UCC was generally well managed during EVD outbreaks. However, they also emphasised that qualified staff (e.g., pharmacists, consultants) were employed to manage this process during well-resourced campaigns. Multi-dose vaccine vials simplified UCC storage and transport, requiring less space than single-dose vials, but one participant (NGO, Central Africa) noted how concerns around vaccine wastage with multi-dose vials affected how vaccination teams interacted with those waiting to be vaccinated. In some instances, vaccination teams would wait for sufficient eligible persons to arrive before opening the vial to limit the risk of vaccine wastage. This approach assumed that community members could wait, sometimes for hours, to receive their vaccination.

> [Vaccination teams] wouldn’t open a vial until [enough] people were there … [community members] would arrive at 8 am and they would still be there at 4 pm, waiting for the other people to turn up. And if [the vaccination team] didn’t get [enough] people they wouldn’t open the vial and they would go away again. (NGO, Central Africa)

#### 4) Vaccination teams

##### Recruiting, training, and resourcing vaccination teams

Participants consistently referred to the importance of recruiting local health workers who have established links to the community, can speak the local language, and can leverage existing rapport to increase uptake, rather than bringing in staff from other regions.

> If we need healthcare people, we try to take the village nurses themselves, we train them in the activity, and they go to meet the community. … So training the local community is very good, not finding the people in the capital and sending them to the village. (Multilateral organisation, West and Central Africa)

However, local recruitment depends on the availability of skilled workers and may place additional workload on already overburdened staff. Seconding staff to vaccination activities can also adversely affect the quality and availability of routine health services. Participants referred to how the outbreak response ‘industry’ during EVD outbreaks drained the local health workforce, with staff leaving routine posts to gain higher salaries in the response.

Recruiting new graduates and recently retired health staff to support large-scale vaccination campaigns was a strategy identified in both the literature and interviews.[29] However, participants were wary of introducing a parallel system that potentially excludes existing, trusted frontline health workers from vaccination campaigns. Some participants also highlighted lengthy recruitment processes as a barrier to bringing on additional staff.

Provision of training for vaccination teams is an essential component of pre-campaign activities,[12] but participants highlighted issues around training quality at the lower levels of the frequently used ‘cascade’ or ‘training-of-trainers’ model. Training deficiencies are often picked up and corrected in the field through supervision, participants outlined, but this is resource intensive. The transition to remote training to comply with COVID-19 physical distancing guidelines has introduced additional barriers to training delivery; many settings have limited access to hardware and stable internet. Participants also raised concerns regarding the quality of training on practical skills through virtual methods.

> Most people couldn’t access the internet and use a virtual platform. … And when you evaluate the performance [of the training], it is very low, because most people couldn’t attend it. (National health agency, East Africa)

Even when in-person training has been possible, physical distancing measures have reduced the capacity of commonly used training facilities, thereby restricting the number of people able to take part.

Inadequate resourcing of vaccination teams and lengthy processes for distributing funding were identified as additional barriers, with participants stating that it was unfair to expect teams to perform without funding.

> [The resources] must be available, and that is very key, available and handed to the people that actually get involved, because if you are going to keep the team in the field for the whole day and they don’t have support or any money to get themselves a drink or something to eat, it is going to have a negative effect. (National health agency, East Africa)

#### 5) Vaccination monitoring and safety surveillance

##### Locally led, integrated vaccination monitoring

The ability to link a person to their vaccination, any subsequent adverse events following immunisation, and any breakthrough infections is an important aspect of vaccination campaign monitoring and can increase community confidence by providing organised, accurate information on the progress of the campaign. However, outside of clinical trials, most interview participants described complex data aggregation processes where indicators held in different datasets were collated and compared. Innovations in electronic case-based surveillance systems, such as the use of QR codes and new modules to track immunisation status and adverse events following immunisation, were mentioned by a small number of participants in relation to COVID-19. Nevertheless, most of these systems still involved a combination of entering data on paper in the field and subsequently inputting the data to electronic systems at higher levels (i.e., district level). Parallel systems and limited standardisation in data collection can severely hamper the analysis and operational use of vaccination coverage, vaccine efficacy, and vaccine safety data.[27]

Participants highlighted how limited access to data held by different organisations negatively affected response efforts. In one example, a participant (NGO, Central Africa) reported that government health staff could not compare newly confirmed EVD cases against the records of those who had been vaccinated due to a lack of access to crucial datasets held by other organisations. Participants described tensions when governments were not sufficiently supported in the ownership, storage, access, and analysis of vaccination data, highlighting the importance of these elements for the success of campaigns.

#### 6) Community engagement and social mobilisation

##### Timing and approach of community engagement

Community engagement and social mobilisation are vital components of a successful vaccination campaign. Participants repeatedly raised the importance of having a clear strategy for engaging with communities, with consistent messaging around the disease, the vaccine, vaccination target groups, and adverse events following immunisation. Some participants linked the success of meningitis A vaccination to the Meningitis Vaccine Project’s detailed communication plan, which was developed, implemented and refined over a few years. Importantly, participants outlined, the objective of the Meningitis Vaccine Project’s communication plan was to build community awareness and expectation for the vaccine well in advance of the campaigns.

> The population was very aware [of meningitis], they were really waiting for this vaccine. When we did the [meningitis A] vaccination campaign … the vaccination centres opened at 8 am, but at 5 am people were already queuing to get the vaccine. (Multilateral organisation, West, Central and East Africa)

While participants recognised that reactive vaccination campaigns face intense time pressures, they insisted that community engagement should precede vaccination campaigns by a minimum of one to two months. This lead-in time is necessary to develop relationships with community members, respond to vaccination queries, and tailor delivery strategies.

Participants stressed that early vaccine acceptance could not be taken as indicative of community sentiment throughout a campaign. Continuous engagement with communities was considered crucial to monitoring community acceptance, responding to concerns around vaccine safety, and explaining the rationale behind target group selection. Reconnecting with communities and providing feedback at the end of the campaign was also deemed essential in enhancing trust and building sustainable relationships that would support future vaccine uptake.

> The feedback to the community leaders is very, very important, because we always seek their permission, their help, their assistance, but we never give them feedback. (Multilateral organisation, West Africa)

Respect, honesty, and meaningful discussion were stressed as principles crucial to building trust:

> There are many things that the communities need to understand. They may not be intellectual … but they are not stupid. They observe and ask relevant questions. So one of the keys is to never hide the truth. Be frank and honest with them, because when they trust you, they trust you forever. But when you lose [their] confidence, it’s difficult. (Multilateral organisation, West and Central Africa)

Communities want to actively participate in discussions about the need for, or benefit of, new vaccines.[44] Providing community members with an opportunity to ask questions and query points about the disease or vaccine is a crucial enabler for vaccination success. Participants outlined how engaged community members often became champions for the campaign.

Participants affirmed that vaccines for well-known and feared diseases, like meningitis A and yellow fever, generally had high community uptake, but social mobilisation was much more difficult if diseases were not known or prioritised by communities. Generating demand for vaccination where communities had other priorities outside the health sector was particularly challenging.

> [Community members] would say “What we need is a bridge, we need a road to be repaired. … When you have those things, bring the [vaccination] request.” (National health agency, West Africa)

Addressing this challenge, participants said, required detailed and focused communication about the disease beyond simple vaccine uptake advocacy; working with communities to ensure they had appropriate information to understand the disease, its severity, and the burden on their communities.[44] Data on disease incidence is critical to these discussions. Some participants highlighted concerns that limited COVID-19 case detection in African countries would negatively impact community perceptions of disease burden and the need for vaccination.

The current limited supply of COVID-19 vaccines presents a paradox for LMICs in terms of community engagement and social mobilisation. While delayed vaccine delivery means countries have more time to plan mobilisation activities, they lack the operational funding required to design and deliver these activities. Further, generating demand in the face of limited supply requires careful balancing of risks; some participants spoke of how they had not yet employed previously used demand generation methods, such as SMS alerts, for COVID-19 for fear of overwhelming their limited vaccine supply.

#### 7) Vaccine confidence

##### Responding to vaccine concerns

Swift, transparent and trusted responses are needed to address vaccine concerns. Participants stated that the longer a rumour circulates, the greater the risk to a vaccination campaign. Influencers — political, religious, and traditional leaders or heads of social groups (e.g., women’s groups, youth groups, or sporting groups) — play an important role in counteracting negative rumours. When rumours were spread by a prominent individual, such as a religious leader, participants described using a similarly influential person to counter those messages. Rumours spread by health professionals were especially difficult to address and were best responded to by other health professionals. National Immunisation Technical Advisory Groups can also play a role in fostering community confidence and dispelling rumours by providing independent, evidence-based information on the disease and the vaccination campaign.

Vaccine hesitancy in the COVID-19 response was perceived as particularly challenging compared to previous campaigns. Participants attributed this to the global nature of the pandemic and the propagation of rumours through social media.

> The problem is that COVID-19 is too publicised, which is already a major obstacle to acceptance. … Unfortunately today, the world cannot control [the information] because everyone has become an expert in COVID-19 vaccination or an expert in COVID-19. (Multilateral organisation, West and Central Africa)

Several participants described how social media tracking and online rumour debunking were incorporated into their COVID-19 response activities. Some countries have partnered with social media and technology companies to provide accurate information through their messaging forums. However, other participants described this as a weaker area of their response, highlighting that further investment was needed to counter the proliferation of misinformation and rumours. While participants described the unique challenge of rumours circulating on social media, they also stressed the importance of continued face-to-face interaction with communities to understand and respond to rumours in-person rather than relying on information distributed online.

Participants noted that rumours around ‘vaccine testing’ were prevalent in both the EVD and COVID-19 responses, but that community risk perceptions were different between the campaigns.

> [During the EVD response] there was lots of talk about being guinea pigs and [people] would say, “yeah, we’re guinea pigs”, but a lot of people went, “actually, I’d rather be a guinea pig and get vaccinated than get Ebola.” … Early on in the COVID-19 outbreak [the population] said, “well, we’re not willing to be guinea pigs for this one because it’s not our problem … we are probably not going to die of COVID-19.” (NGO, Central Africa)

Participants spoke of how they explained the vaccine regulatory and approval processes to communities, highlighting that COVID-19 vaccine trials had already been conducted in other countries outside of Africa. However, changing community perceptions was considered difficult due to misinformation and the absence of high disease incidence or severity that might increase the prioritisation of COVID-19 vaccination within communities.

## DISCUSSION

This research took place as the first doses of COVID-19 vaccines provided through COVAX arrived in LMICs in Africa and South America. We reviewed the literature and interviewed technical experts with extensive experience in epidemic vaccination campaigns to develop recommendations for the rollout of COVID-19 vaccines in LMICs. Table 4 presents a set of recommendations. Key recommendations include: prioritising time, funding and workforce for community engagement; identifying effective training strategies to upskill vaccination teams; streamlining response coordination and vaccination monitoring functions; and exploring opportunities for health service integration.

**TABLE 4:**
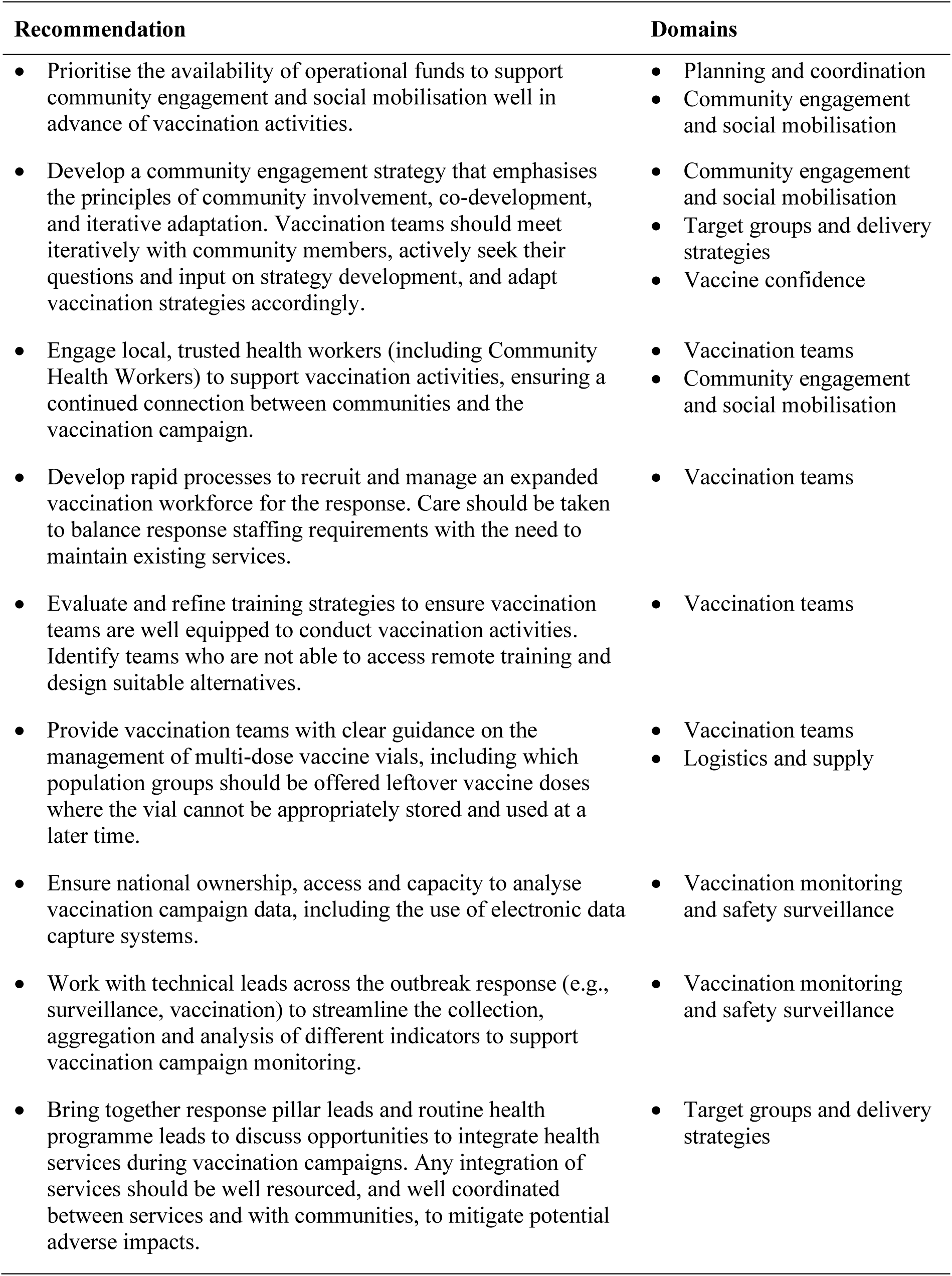
Recommendations for the implementation of COVID-19 vaccines in LMICs, based on lessons from meningitis A, yellow fever and Ebola virus disease vaccination campaigns.

To date, gaps in operational funding for COVID-19 vaccination campaigns in LMICs have prevented some vaccination teams from conducting crucial community engagement activities. Mobilising communities in the face of limited COVID-19 vaccine supply is a difficult balance. Nevertheless, while waiting for sufficient vaccines to be delivered, countries can involve communities in the proposed rollout, listen to their concerns and devise solutions. It is crucial that countries prioritise operational funds to support vaccination teams to effectively conduct these preparatory activities.

There are several advantages to employing local health workers in vaccination campaigns. Building on existing trust and rapport, local health workers can engage with communities, discuss key information about the campaign, monitor vaccine acceptance levels and respond to rumours as they arise. In order to employ local health workers that are not yet supporting the COVID-19 response, countries must identify mechanisms to rapidly recruit and train them. Evidence around effective training strategies for health workers in LMICs during epidemics remains limited.[49, 50] While the ‘cascade’ or ‘training-of-trainers’ model is seen as economical and rapidly scalable,[51] participants highlighted issues of quality as information is transmitted to lower levels. Considering the key role health workers play in mobilising communities and delivering vaccinations, countries must evaluate and refine training strategies to effectively upskill their vaccination teams for COVID-19.

Co-administration of COVID-19 with other vaccines is not currently recommended by the WHO Strategic Advisory Group of Experts on Immunization,[52] preventing potential efficiency gains through integration. While integration can reduce time and cost burdens, it introduces challenges around coordination, reporting, and staff capacity.[53] Further, COVID-19 vaccine hesitancy may affect the uptake of other vaccines. The differentiation between COVID-19 and other vaccines has perpetuated the belief that some vaccines are safe while others are not. Without a concerted effort to change community perceptions around COVID-19 vaccines, integrating other campaigns with COVID-19 vaccination is unlikely to yield high uptake. Integrating other community-prioritised health services (e.g., antenatal care) may be a more effective strategy. Careful planning with other health programmes and communities is needed to identify and mitigate the potential effect of COVID-19 vaccine hesitancy on other services. Considering the significant impact of COVID-19 on access to routine health services in LMICs,[54, 55] the integration of services should be discussed and pursued where appropriate.

Most COVID-19 vaccines currently under Emergency Use Listing (EUL) by WHO are packaged in multi-dose vials.[21, 56] Multi-dose vaccine vials are widely used in LMICs as they are cheaper and require less storage space.[57] However, health workers’ reluctance to open multi-dose vials for fear of vaccine wastage was identified in this research and has also been reported for routine immunisation programmes in LMICs.[21, 56] The limited supply of COVID-19 vaccines is likely to increase pressure on vaccination teams to minimise wastage, which may be complicated by the strict prioritisation of target groups and instances where sufficient eligible persons do not present for vaccination. To minimise wastage and reduce the risk of loss-to-follow up, vaccination teams should be given clear guidelines for administering the remaining doses in multi-dose vials to persons outside of priority groups.

Incident management systems provide direction and coordination across response activities, including vaccination. However, the unprecedented scale and extended duration of the COVID-19 pandemic has placed enormous pressure on key personnel within response structures. The incident management system model is labour intensive.[58, 59] While there has been a global recognition of frontline worker burnout, the effect of a protracted pandemic on key personnel in countries’ incident management systems should also be considered.[60] Countries need to explore ways to streamline coordination processes and upskill additional staff to fulfil IMS functions to ensure continuity in the response.

Due to the geographical distribution of the selected diseases and the distribution of the research team’s professional networks, this research draws primarily from vaccine implementation experiences in LMICs in West, Central and East Africa. The findings may not be generalisable to other geographic areas, which are likely to have their own set of challenges in implementing vaccines. In addition, several identified technical experts could not participate due to competing priorities, including the COVID-19 response and the EVD outbreaks in the first half of 2021.

## CONCLUSION

These recommendations rely on LMICs having sufficient vaccine supply to conduct vaccination campaigns for COVID-19, which has not been the case in most settings. We implore the global community to prioritise COVID-19 vaccine supply for LMICs. As vaccine supplies increase, we encourage researchers to support countries in monitoring and documenting their COVID-19 vaccination campaigns to understand real-time responses to challenges and strengthen evidence around best practices during outbreak-related vaccination campaigns in low-resource settings.

## Data Availability

The qualitative data generated through this study are not suitable for sharing beyond that contained within the manuscript in order to protect the anonymity of participants. Further information can be obtained from the corresponding author.

## Acknowledgements

The authors would like to thank the technical experts who took time out of their busy schedules in the midst of the COVID-19 pandemic and concurrent outbreaks to give interviews and who generously shared their experiences, knowledge and insights to support this research.

## Contributors

All authors provided input into the conceptualisation of the study and its design. The interviews and qualitative analysis were conducted by J.C and R.W with support from D.K. The literature review was conducted by D.K with support from K.N and J.C. The manuscript was prepared by J.C and D.K. All authors provided input into the manuscript and are guarantors of the study.

## Patient consent

Not required.

## Ethics approval

The study was approved by the London School of Hygiene & Tropical Medicine Research Ethics Committee (ref: 24044).

## Provenance and peer review

Not commissioned; externally peer reviewed.

## Data sharing statement

The qualitative data generated through this study are not suitable for sharing beyond that contained within the manuscript in order to protect participants’ anonymity. Further information can be obtained from the corresponding author.

