## Appendix 1 for "“Learn from the lessons and don’t forget them”: Identifying transferable lessons for COVID-19 from meningitis A, yellow fever, and Ebola virus disease vaccination campaigns"

### APPENDIX 1: Literature review search terms

***MEDLINE***

| |
|---|
| 1. meningococcal vaccines/ or ebola vaccines/ or yellow fever vaccine/ |
| 2. ("ebola" or "yellow fever" or "meningitis a" or "meningococcal a" or "meningococcal meningitis").ab,ti,kw. |
| 3. meningitis, meningococcal/ or hemorrhagic fever, ebola/ or yellow fever/ |
| 4. 2 or 3 |
| 5. ("immuni?ation" or "vaccin*" or "ervebo" or "17d" or "MenAfriVac").ab,ti,kw. |
| 6. 1 or 5 |
| 7. 4 and 6 |
| 8. Vaccines/ad, lj, st, sd [Administration & Dosage, Legislation & Jurisprudence, Standards, Supply & Distribution] |
| 9. Immunization Programs/lj, mt, og, st, sd [Legislation & Jurisprudence, Methods, Organization & Administration, Standards, Supply & Distribution] |
| 10. implementation science/ |
| 11. "Delivery of Health Care"/ |
| 12. 8 or 9 or 10 or 11 |
| 13. 7 and 12 |

***GLOBAL HEALTH***

| |
|---|
| 1. ("immuni?ation" or "vaccin*" or "ervebo" or "17d" or "MenAfriVac").ab,ti,sh. |
| 2. ("ebola" or "yellow fever" or "meningitis a" or "meningococcal a" or "meningococcal meningitis").ab,ti,sh. |
| 3. (("immunisation" or "vaccin*") adj3 ("uptake" or "strateg*" or "feasibility" or "program*" or "campaign" or "delivery" or "roll?out")).ab,ti,sh. |
| 4. 1 and 2 and 3 |

***EMBASE***

| |
|---|
| 1. meningococcal vaccines/ or ebola vaccines/ or yellow fever vaccine/ |
| 2. ("ebola" or "yellow fever" or "meningitis a" or "meningococcal a" or "meningococcal meningitis").ab,ti,kw. |
| 3. meningitis, meningococcal/ or hemorrhagic fever, ebola/ or yellow fever/ |
| 4. 2 or 3 |
| 5. ("immuni?ation" or "vaccin*" or "ervebo" or "17d" or "MenAfriVac").ab,ti,kw. |
| 6. 1 or 5 |
| 7. 4 and 6 |
| 8. vaccination coverage/ or vaccination refusal/ |
| 9. implementation science/ |
| 10. (("immunisation" or "vaccin*") adj2 ("uptake" or "strateg*" or "feasibility" or "program*" or "campaign" or "delivery" or "roll?out")).ab,ti,kw. |
| 11. 8 or 9 or 10 |
| 12. 7 and 11 |
